# SARS-CoV-2 Delta variant of concern in Brazil - multiple introductions, communitary transmission, and early signs of local evolution

**DOI:** 10.1101/2021.09.15.21262846

**Authors:** José Patané, Vincent Viala, Loyze Lima, Antonio Martins, Claudia Barros, Elaine Marqueze, Jardelina Bernardino, Debora Moretti, Svetoslav Slavov, Rafael Bezerra, Evandra Rodrigues, Elaine Santos, Ricardo Brassaloti, Raquel Cassano, Pilar Mariani, João Kitajima, Bibiana Santos, Patricia Assato, Felipe Costa, Mirele Poleti, Jessika Lesbon, Elisangela Chicaroni, Cecilia Banho, Lívia Sacchetto, Marília Moraes, Melissa Palmieri, Fabiana Vilanova, Maiara Martininghi, Rejane Grotto, Jayme Souza-Neto, Marta Giovanetti, Luiz Alcantara, Maurício Nogueira, Heidge Fukumasu, Luiz Coutinho, Rodrigo Calado, Simone Kashima, Raul Neto, Dimas Covas, Carolina Elias, Sandra Coccuzzo

## Abstract

The dissemination of the Delta VOC in Brazil is still unclear, despite the frequent reports of isolated cases from different Brazilian states. In this report we characterize the dissemination of the Delta VOC in Brazil and where the introductions of this lineage fall within the global Delta phylogeny. We also examined the mutational profile of the largest clade within the Brazilian Delta VOCs, with a focus on samples which were obtained in the State of São Paulo, and especially in the city of São Paulo, the largest metropolis of South America, and a national and international transportation hub.

## 1. Introduction

Brazil has been a continuous source of new Covid-19 variants since 2020 [1] especially with the emergence of P.1 (Gamma), a variant of concern (VOC) which has spread throughout the whole country since January 2021, accounting for more than 90% of the cases weekly [2]. At the same time, Brazil is a world exporter and importer of SARS-CoV-2 lineages [1]. This is especially alarming considering the Delta VOC, which emerged in India in early September, 2020 and led to a massive death toll locally. The Delta VOC is now spread worldwide, contributing to a relevant rise in hospitalizations and deaths. As an illustrative example of its impact, in the UK it surpassed the previously dominant VOC, the B.1.1.7 (Alpha) lineage (source: outbreak.info). The dissemination of the Delta VOC in Brazil, first detected by Lamarca et al. [3], is still unclear, despite the frequent reports of isolated cases from different Brazilian states like São Paulo (SP), Rio de Janeiro (RJ), Minas Gerais (MG), Rio Grande do Sul (RS), Tocantins (TO), Paraná (PR), Goiás (GO), and Maranhão (MA). In this report we characterize the dissemination of the Delta VOC in Brazil and where the introductions of this lineage fall within the global Delta phylogeny. We also performed characterization of the mutational profile of the largest clade within the Brazilian Delta VOCs, with a focus on samples which were obtained in the State of São Paulo and especially in the city of São Paulo, the largest metropolis of South America, and a national and international transportation hub.

## 2. Materials and Methods

From all SARS-CoV-2 positive samples belonging to the Laboratory Platform for Coronavirus Diagnosis, established by the Butantan Institute, we randomly select and sequence around 7-10% from each epidemiological week (epiweek). Viral genotyping was performed only on samples with cycle threshold (Ct) values up to 35. Briefly, RNA extraction was performed with the Extracta kit AN viral (Loccus) in an automated extractor (Extracta 32, Loccus) following the manufacturer’s guidelines. SARS-CoV-2 molecular diagnosis was carried out using the GeneFinderTM COVID19 Plus RealAmp kit (Osang Healthcare Co. Ltd.) in all laboratories comprising the diagnostic platform, which reduces variations related to Ct values.

SARS-CoV-2 genomic libraries were generated using the COVIDSeq kit (Illumina, San Diego, CA) following the manufacturer’s specifications. The normalized sample libraries were sequenced on a Illumina MiSeq instrument (Kit v2, 2×300) (Illumina, San Diego, CA, USA). The obtained Delta sequences were of high quality, with mean read number above 200,000, mean depth above 800, and coverage above 99%. Reference-assembly following a modified pipeline after the COVIDSeq protocol (https://artic.network/ncov-2019) was used. Assembled genomes were deposited in GISAID (accessions provided in Suppl. Mat. Table S1).

In order to contextualize the evolution of Brazilian Delta genomes internationally, we assembled a global and clade-representative vetted dataset of 4.000 sequences available from GISAID, spanning a plethora of lineages (including Delta). Subsequently we blasted (Blastn) each Brazilian Delta genome separately and collected only the 1,000 closest genomes of each Brazilian sample, using the whole set of more than 170,000 Delta genomes available on GISAID (as of July 19^th^, 2019) as the Blast database. Duplicate entries were then removed. Finally, we included eventual Brazilian Deltas that had been left out of either of those two data sets (either the global representative, or the Blastn-derived). A total of 11,742 genomes were included for phylogenetic inference, including 98 Brazilian Delta VOC sequences, out of which 13 were obtained in this study from the city of São Paulo, spanning epiweeks 25 to 27. Additionally, a larger set of Delta VOC genomes from São Paulo state was established up to epiweek 29 to refine Delta’s growth curve as well as to show this variant has been disseminated along the State.

A maximum-likelihood (ML) tree was obtained [4], and only the Delta clade plus its neighbor clade (B.1.6171) were kept for subsequent analyses, for a total of 8,214 samples. Tip-dating was employed to assess divergence times [5]. Branches with approximate likelihood support (aLRT) lower than 0.8 were collapsed. Protein-changing substitutions were assessed by checking individual gene alignments (https://github.com/neherlab/nextalign).

## 3. Results and Discussion

According to our analyses, at least 10 independent introductions have occurred until performing the present study in Brazil (clades I-X in Fig. 1). Some of them were imports related to either Australia (clades III and V; Figs. 1 and 2) or USA (clade III), while others are more related to UK samples (clades VI and VIII; Fig. 2 - also clades VII and IX, not shown). Others clustered within a large polytomy with numerous international samples hence precluding accurate source inference (e.g., clade I; Fig. 2). Four clades included three or more samples (branch support >= 0.8): clade I (3 from Parana State), clade III (71 tips, mostly from Rio de Janeiro, but also 4 obtained from São Paulo, 1 from Rio Grande do Sul, and 1 from Tocantins), clade VI (6 from the State of Maranhão), and clade VIII containing three samples from Goiás State. We also mention a cluster of three SP and and two PR sequences in cluster V, but these do not form an exclusive clade, being also closely related to Australian samples.

**Figure 1.**
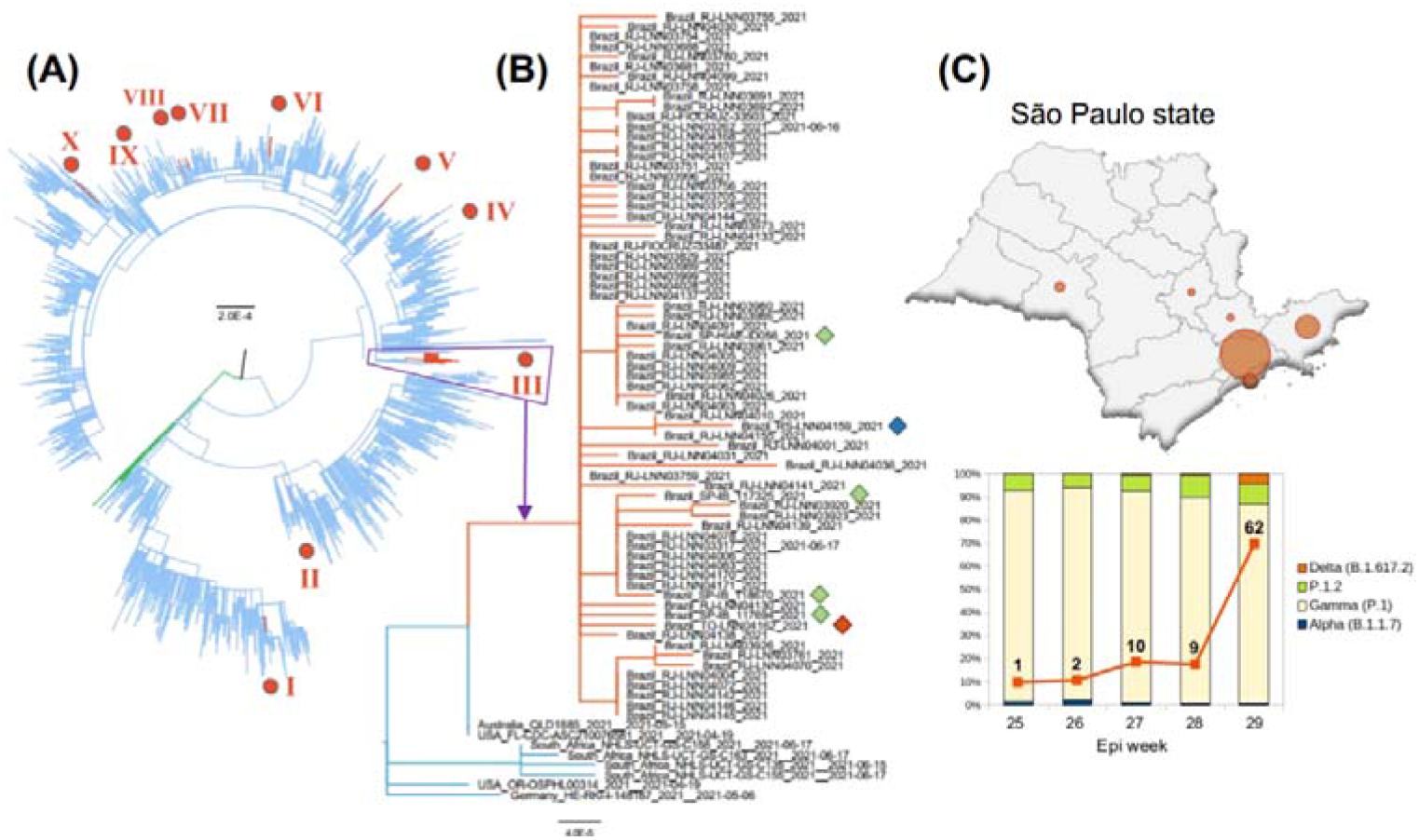
Aspects of Delta evolution in Brazil. (A) ML tree with 8,214 samples showing phylogeny of this variant (blue), with lineage B.1.617.1 (green) as outgroup (other lineages basal relative to the above are not shown). Brazilian Delta samples (epiweeks 25-27) are indicated in red with roman numerals. All branches < 0.8 support were collapsed. (B) Clade III, the largest clade, with samples from RJ, SP (green diamonds), RS (blue), and TO (red). (C) top: SP state health departments with presence of Delta samples proportional to circle size (epiweeks 25-29); bottom: variation in Delta frequency in SP from epiweeks 25-29 (absolute and relative to lineages most commonly found).

**Figure 2.**
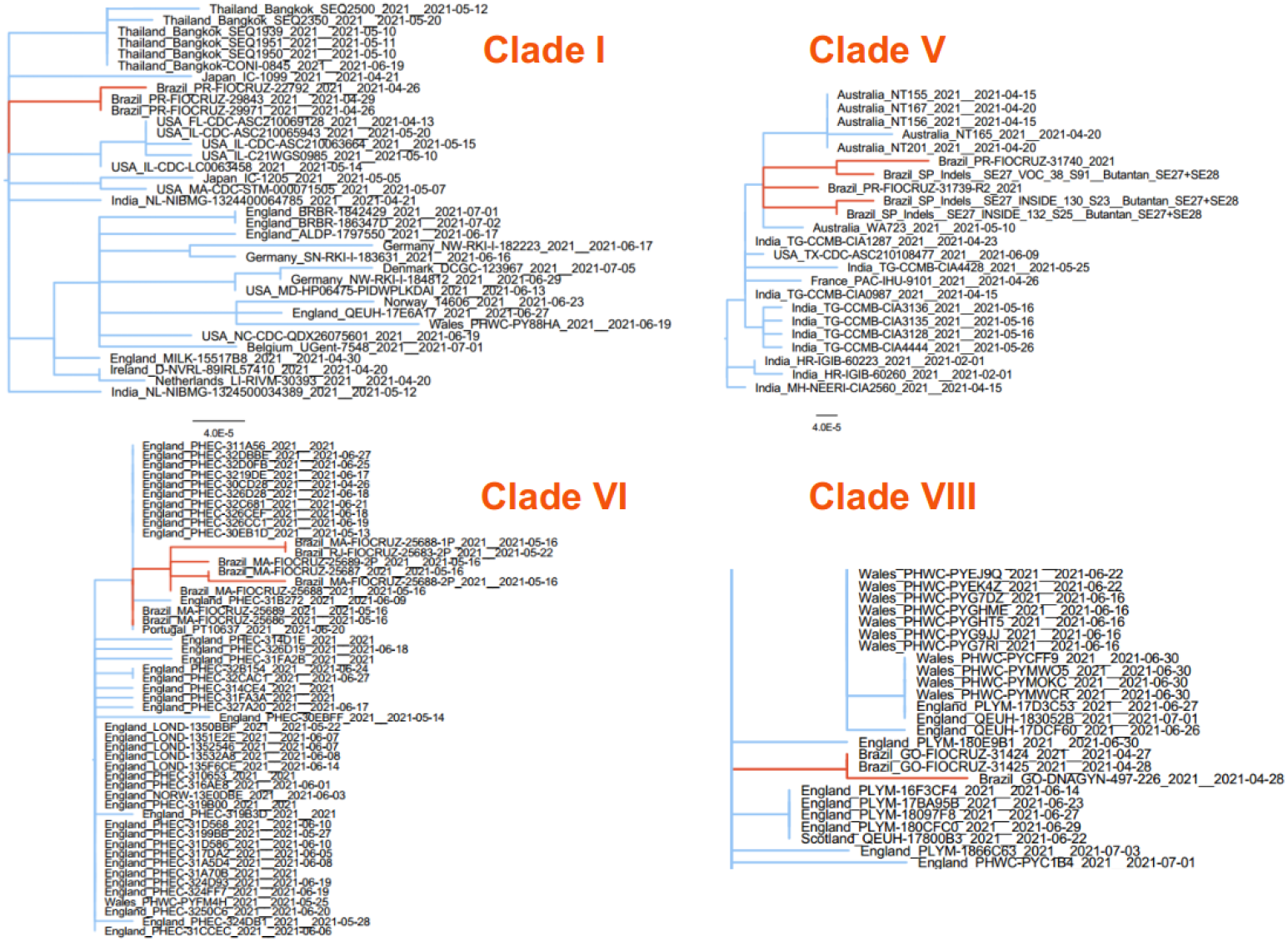
Clades encompassing at least three Brazilian Delta genomes.

We focus on the largest clade (III) embracing Rio de Janeiro, São Paulo, Tocantins and Rio Grande do Sul samples (Fig. 1), which was also the clade with largest internal branch across Brazilian samples, denoting early signs of local evolution within the country. This clade encompasses 71 genomes (mostly from RJ), further showing its importance in terms of surveillance concern. The mutational profile revealed three mutations exclusive to this clade, ORF1a: T4087I (ORF1a), and the remaining two being synonymous. The T4087I mutation is related to a change of a hydrophilic amino acid (Thr) to hydrophobic (Ile). ORF1a does not embrace any structural proteins, but it may be possible that such a mutation affects the replication rate (hence viral load), which can in turn impact infection rates; more studies are needed to explore such putative effects of this substitution.

The first reported cases of Delta VOC infections in Brazil are associated with a cargo ship departed from Malaysia on March 27th, and with connection in South Africa that had arrived in Brazil by May 14th, carrying over 20 crewmen, six of which positive for Delta. Initially communitary transmission had been ruled out, but our results indicate otherwise (clade VI, Fig. 2). Regarding the largest clade with communitary transmission (clade III, Fig. 1), its most recent common ancestor (MRCA) is from early June 2021 according to dating inference, suggesting a recent expansion unrelated to MA cases.

We detected at least four independent communitary transmission chains state-wise (one in RJ, one in GO, another in MA, and also in PR; Figs. 1 and 2). At the same time, some samples from the same state belong to different clades (e.g., SP samples in clades III and V; RJ in clades III and VI; PR in clades I and V), evincing the multiple introductions mentioned above.

Sao Paulo, the largest city of the American continents, has samples mainly from its northern districts (Suppl. Mat. Table S1). The first collected sample in the city of Sao Paulo (from late June) is from the southeastern region of the Great São Paulo though, where it probably arrived via the Taubate Regional Health Department from RJ. Presently (epiweek #29), 84 samples have been found in six distinct Regional Health Departments along Sao Paulo state (Fig. 1C), indicating the Delta VOC is gaining momentum within the state, raising concern regarding its potential impact on hospitalizations, deaths, and vaccine escape (as already observed in many countries).

## 4. Conclusions

The SARS-CoV-2 Delta variant (B.1.617.2) is now disseminated worldwide. Besides the UK (where it surpassed the previously dominant Alpha VOC), a more comparable scenario to Brazil (in populational, geographic, economic and cultural terms) is Mexico, where the Gamma variant had been pervasive, yet the Delta VOC has been able surpass it in distribution by mid-June 2021 (source: outbreak.info). The increase in Brazilian cases reported here may serve as an alert, especially given the SARS-CoV-2 impact on the country’s health care system, which has been severely affected by both COVID-19 hospitalizations and deaths. In support of this is the fact that Brazil is a world leader of COVID-19 morbidity and mortality (554, 626 deaths by July, 30th, surpassed only by the USA). In our study, we expanded on the dataset used by the report by Lamarca et al. [3], allowing a more in-depth assessment of the origin and spread of this VOC in Brazil.

Our analyses showed at least 10 different Delta VOC introductions and its spread over eight Brazilian states in the first semester of 2021, which was not unexpected given the currently limited contact tracing surveillance protocols in Brazil and the vast national territory. Four apparently unrelated transmission chains (here defined as a supported clades encompassing at least three samples) were identified (in the states of São Paulo, Paraná, Goiás and Maranhão), indicating that the presence and transmission of the Delta VOC has already been established in Brazil. Finally, the locally fixed mutation (ORF1ab: T4087I) which changes a hydrophilic amino acid for a hydrophobic one, was also observed in the report by Lamarca et al. [3], raising the concern aimed at this lineage.

Even though preliminary analyses indicate that SARS-CoV-2 vaccines used nationwide may offer protection against the Delta VOC [6], the introduction of any VOC in a country demands significant attention. In this respect, activities related to evaluation of Delta VOC evolution and dissemination with the use of continuous SARS-CoV-2 molecular vigilance in Brazil must be widely implemented thus reducing the possible impact of this VOC on both healthcare system and vaccination process.

## Data Availability

I am making available all data referred to in the manuscript in the form of tables (either regular os supplementary).

## Author Contributions

Conceptualization, J.P., V.V., C.S., S.K., S.C., D.C.; methodology, J.P., C.S., S.K., S.C.; software, J.P., R.S., V.V.; validation, J.P., V.V., R.S., C.S. and S.K.; formal analysis, J.P., V.V., C.S., S.K., S.C., D.C.; investigation, J.P., V.V., C.S., S.K., S.C., S.S., D.C.; resources, R.N., D.C., S.C., C.S., S.K.; wet lab, V.V., E.R., E.S., L.L., J.B., C.B., E.M., R.B., R.C., P.M., J.K., B.S., P.A., F.C., M.P., J.L., C.S., C.B., L.S., M.G., R.C., M.M., M.P., F.S.; data curation, A.M., S.C., C.S., S.K., V.V., J.P.; writing—original draft preparation, J.P., V.V., S.S., R.S., A.M., R.G., J.N., L.A., M.N., H.F., L.C., S.K., C.S., S.C.; visualization, J.P., V.V., C.S., S.K., S.C.; supervision, C.S., S.K., S.C.; project administration, D.C., S.C.; C.S., S.K.; funding acquisition, D.M., R.G., J.N., L.A., M.N., H.F., L.C., R.C., S.K., C.S., S.C, J.K., D.C. All authors have read and agreed to the published version of the manuscript.

## Institutional Review Board Statement

The study was conducted according to the guidelines of the Declaration of Helsinki, and approved by the Institutional Ethics Committee of the Faculty of Medicine of Ribeirão Preto, University of Sao Paulo (CAAE: 50367721.7.1001.5440).

## Informed Consent Statement

Patient consent was waived once the diagnostic test had already been performed and the result has been communicated to the patients. Viral RNA was used and the results obtained will not compromise private information, neither bring any new clinical/therapeutic outcome for the patient.

## Acknowledgments

We thank all contributors from GISAID. We also thank Gabriela Ribeiro and Glaucia Borges for their help with the Instituto Butantan’s local database repository.

